# “Association between COVID-19 vaccination, infection, and risk of Guillain-Barre syndrome, Bell’s palsy, encephalomyelitis and transverse myelitis: a population-based cohort and self-controlled case series analysis”

**DOI:** 10.1101/2021.09.08.21263276

**Authors:** Xintong Li, Eugenia Martinez-Hernandez, Elena Roel Herranz, Antonella Delmestri, Talita Duarte-Salles, Victoria Strauss, Edward Burn, Daniel Prieto-Alhambra

**Author notes:** joint first authors (Xintong Li, Eugenia Martinez-Hernandez). joint senior authors (Edward Burn, Daniel Prieto-Alhambra). **CORRESPONDING AUTHOR** Dr Victoria Strauss, Lead statistician, Centre for Statistics in Medicine, NDORMS, University of Oxford, Botnar Research Centre, Windmill Road, OX37LD, Oxford, United Kingdom.

## Abstract

**OBJECTIVE:** We aimed to study the association between COVID-19 vaccines, SARS-CoV-2 infection, and the risk of immune-mediated neurological events.

**METHODS:** *Design:* Population-based historical rate comparison study and self-controlled case series (SCCS) analysis.

*Setting:* Primary care records from the United Kingdom.

*Participants:* Individuals who received the first dose of ChAdOx1 or BNT162b2 between 8 December 2020 and 6 March 2021. A cohort with a first positive RT-PCR test for SARS-CoV-2 between 1 September 2020 and 28 February 2021 was used for comparison.

*Main outcome measures:* Outcomes included Guillain-Barre syndrome (GBS), Bell’s palsy, encephalomyelitis, and transverse myelitis. Incidence rates were estimated in the 28 days post first-dose vaccine, 90 days post-COVID-19, and between 2017 to 2019 for the general population cohort for background rates. Indirectly standardised incidence ratios (SIRs) were estimated. Adjusted incidence rate ratios (IRR) were estimated from the SCCS when sufficient statistical power was reached.

*Results:* We included 1,868,767 ChAdOx1 and 1,661,139 BNT162b2 vaccinees; 299,311 people infected with COVID-19; and 2,290,537 from the general population. SIRs for GBS were 1.91 [95% CI: 0.86 to 4.26] after ChAdOx1, 1.29 [0.49 to 3.45] after BNT162b2, and 5.20 [1.95 to 13.85] after COVID-19. In the same cohorts, SIRs for Bell’s palsy were 1.34 [1.05 to 1.72], 1.15 [0.88 to 1.50], and 1.23 [0.80 to 1.89], and for encephalomyelitis 1.62 [0.61 to 4.31], 0.86 [0.22 to 3.46], and 11.05 [5.27 to 23.17], respectively. Transverse myelitis was too rare to analyse (n<5 in all cohorts). SCCS analysis was only conducted for Bell’s palsy due to limited statistical power. We found no association between either vaccine and Bell’s palsy, with an IRR of 1.10 [0.81 to 1.46] and 1.15 [0.87 to 1.49] for BNT162b2 and ChAdOx1, respectively.

*Conclusions:* We found no consistent association between either vaccine and any of the studied neuroimmune adverse events studied. Conversely, we found a 5-fold increase in risk of GBS and an 11-fold of encephalomyelitis following COVID-19.

## INTRODUCTION

As of 3 September 2021, the COVID-19 pandemic has caused more than 4.5 million deaths worldwide. After rapid development, 5.2 billion vaccine doses against SARS-CoV-2 have been administered through national vaccination programs.[1] The AstraZeneca (ChAdOx1) and Pfitzer-BioNTech (BNT162b2) vaccines were the first to be approved for use in the United Kingdom in December 2020. They have shown high effectiveness in preventing severe COVID-19 in clinical trials and real-world settings.[2–4] Over 91 million doses of the vaccine have been administered in the UK, and 43 million people have been fully vaccinated, which is 79% of the population.[5] As immunisation programs continue to advance, safety monitoring is crucial to assess adverse events and identify potential risks.

Immune-mediated neurological disorders among individuals vaccinated with adenovirus-based vaccines against SARS-CoV-2 have been identified. Up to 25 August, the Medicines and Healthcare products Regulatory Agency (MHRA) has received 393 reports of Guillain-Barré syndrome (GBS) and 23 reports of Miller-Fisher syndrome (a variant of this disease) after ChAdOx1, 44reports of GBS after BNT162b2, and 3 reports of GBS after the Moderna (mRNA-1273) vaccine.[6] Instances of these and other neuro-immune events also have been reported elsewhere. Reports after ChAdOx1 include seven cases of GBS in India,[7] two in the UK,[8,9] one in each of in France, [10] Italy,[11] and Malta,[12] and nine cases of bifacial weakness with paresthesias, a variant of GBS, in the UK,[13,14] and one in Italy,[15] and two cases of sensory GBS in Korea.[16] Three cases of acute transverse myelitis were reported during the clinical trials for ChAdOx1,[17,18] two in case studies,[19,20] and one report of longitudinal extensive transverse myelitis after ChAdOx1 vaccination reported in a case study.[21] Reports after the BNT162b2 vaccine include one case of GBS in each of the United States (US) and Qatar,[22,23] two in Italy,[24,25] and seven among a cohort of 3,89 million vaccinees in Mexico,[26] and one case of acute disseminated encephalomyelitis.[27] Data from the BNT162b2 and mRNA-1273vaccines trials also suggested a possible association with Bell’s palsy, with seven cases in the active arms compared with one case in the placebo arms of the trials, and six subsequent reports.[28–33] The clinical trial of the Johnson & Johnson viral vector vaccine Ad26.COV2.S reported one case of GBS in each of the placebo and active trial arms 10 days after the injection.[34,35] There were 2 reports of the GBS variant with bifacial weakness in the US,[26,36] and a warning of the US Food and Drug Administration about the Ad26.COV2.S vaccine and rare but serious cases of GBS.[37] One case of Bell’s palsy and one of transverse myelitis with Bell’s palsy reported after Ad26.COV2.S vaccination

Although these reports do not imply causality, the temporal association between vaccination and neurological symptoms warrants robust post-vaccination surveillance. Large-scale epidemiologic studies are required to determine whether COVID-19 vaccination increases the risks above background rates in the general population.

We leveraged large routinely collected datasets including millions of vaccinated people in the UK to study the potential association between COVID-19 vaccination and the risk of developing GBS, Bell’s palsy, encephalomyelitis, and transverse myelitis. To help contextualise our results, we also studied the associations between infection with SARS-CoV-2 and the risk of the studied immune-mediated neurological outcomes.

## METHODS

### Data sources

Data were obtained from the UK Clinical Practice Research Datalink (CPRD) GOLD and the CPRD Aurum. CPRD GOLD and CPRD Aurum contain routinely collected data from primary care practices in the UK,[38,39] representing 5% and 20% of the current UK population, respectively.[40,41] Both databases have been mapped to the Observational Medical Outcomes Partnership (OMOP) common data model for curation and analysis.[42]

### Study participants

The populations of interest were individuals who had received their first dose of a COVID-19 vaccine and people infected with SARS-CoV-2, as identified from CPRD AURUM. Using the CPRD GOLD data, we created a background population cohort. The two CPRD databases were used because the CPRD GOLD data was only available until mid-2020..

Two mutually exclusive cohorts were constructed of people vaccinated with first-dose ChAdOx1 or BNT162b2 between 8 December 2020 and 6 March 2021. The date of the first vaccine dose administration was used as the index date. The SARS-Cov-2-infected cohort included people with a first positive RT-PCR test between 1 September 2020 and 28 February 2021, with the test date used as the index date. The background population cohort were people with a primary care contact recorded between 1 January 2017 and 31 December 2019. The index date for the general population cohort was the first visit date during this period.

All participants were required to be aged 30 years or older and to have at least 365 days of visibility in the data before the index date. For each cohort, participants were censored at the first of a health outcome of interest, end of data availability (14 March 2021), or leaving the database (death or migration).

### Events of interest

The events of interest were four immune-mediated neurological disorders prespecified as potential adverse events of special interest for COVID-19 vaccine safety: Guillain-Barré syndrome, Bell’s palsy, encephalomyelitis, and transverse myelitis.[43] Events were identified using previously validated clinical codes from electronic health records.[43] Details of the Systematized Nomenclature of Medicine (SNOMED) codes used to define the outcomes are available in Appendix Table 1.

### Study design

We used the historical rate comparison method. Incidence rates of each outcome in the vaccinated cohorts and SARS-Cov-2-infected cohort were used as “observed” rates and compared with the “expected” background incidence rates estimated from the general population cohort. For the vaccinated cohorts, we estimated the rates during the 1 to 28 days after the first-dose vaccination (day 0). A 90-days post-test period was used for the SARS-Cov-2-infected cohort.

A self-controlled case series (SCCS) method was then applied where possible to minimise confounding. SCCS is an established method for vaccine safety surveillance, which includes only individuals experiencing both exposure and outcome, and participants act as controls for themselves, therefore eliminating time-fixed confounding.[44] Within-person comparisons of event rates were made between the pre-vaccination (baseline) and the post-vaccine exposure period. Similar to the historical rate comparison design, we defined the exposure window as the 1 to 28 days after first-dose vaccination (day 0). The study period of the SCCS analysis is from 1st January 2017 to the end of data availability. (Figure 2)

**Figure 1.**
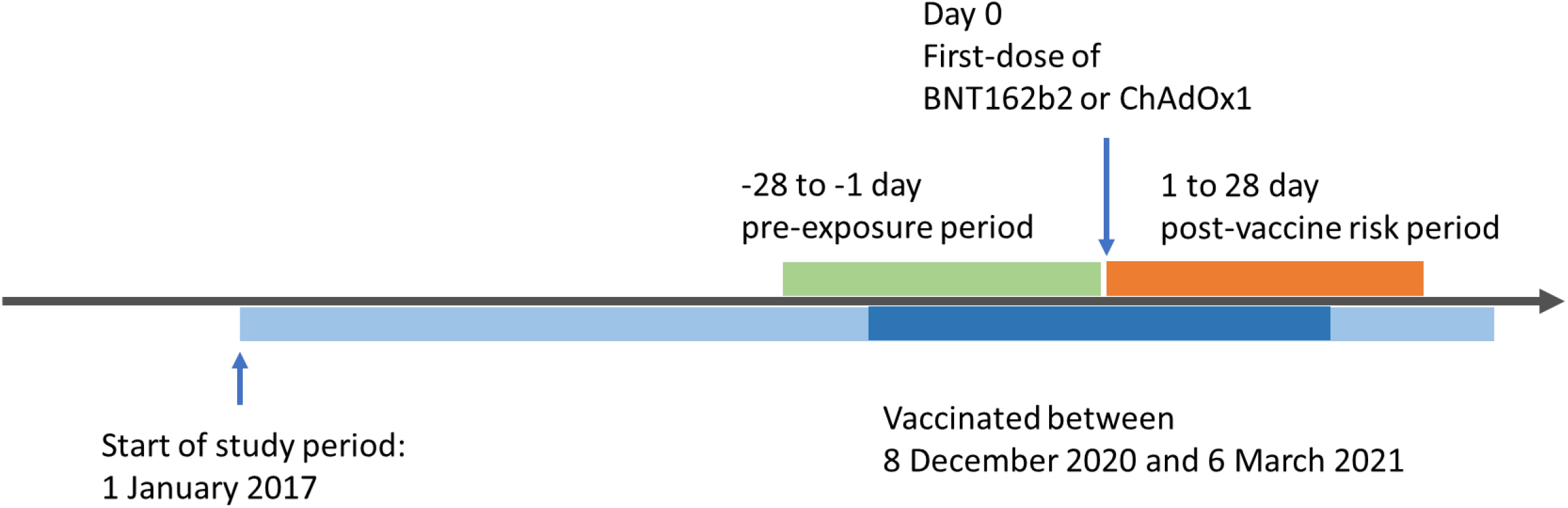
Self-controlled case series analysis design.

**Figure 2.**
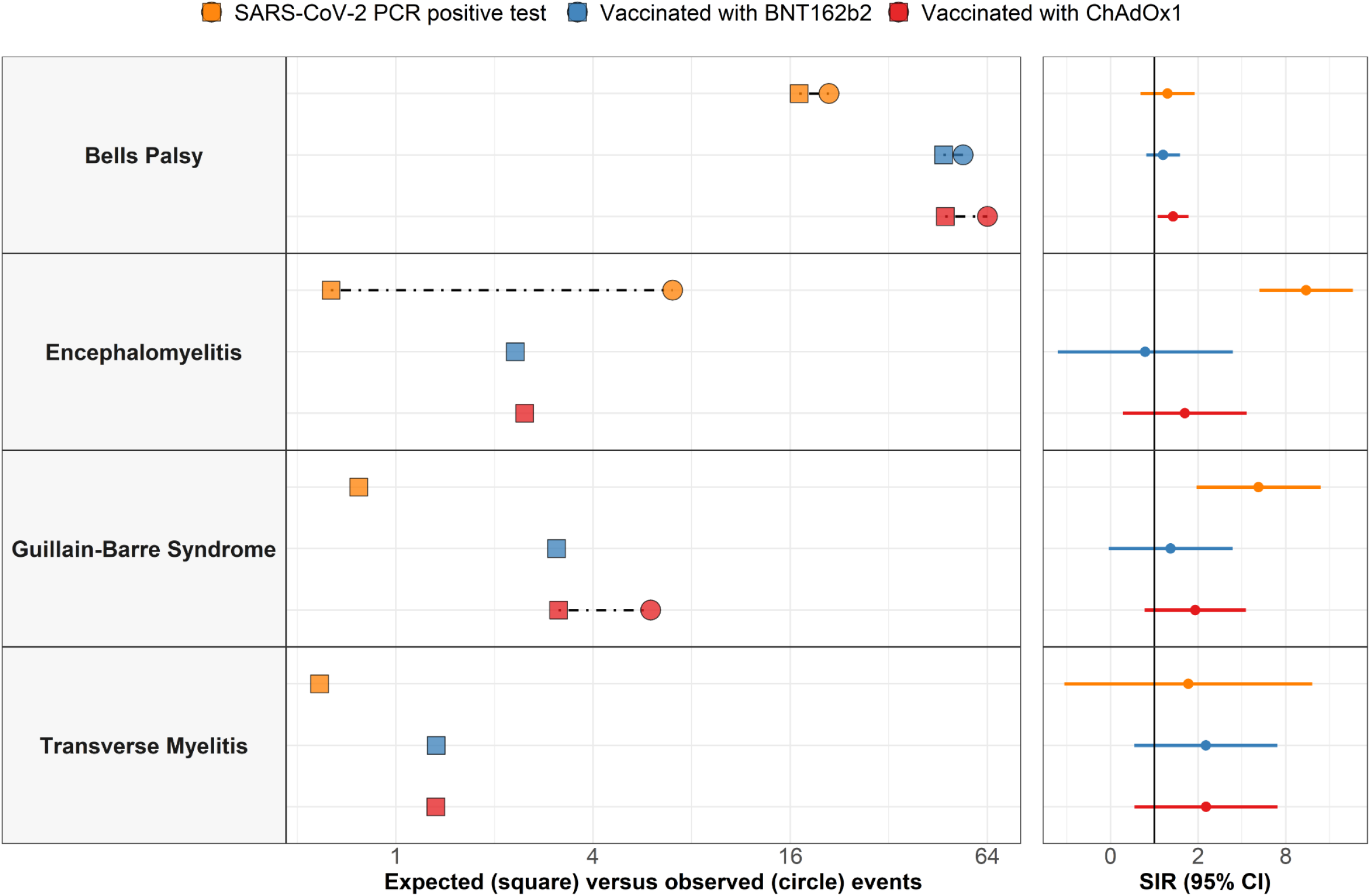
Expected vs observed rates of neuro-immune adverse events of special interest and standardised incidence ratios (SIRs)

We also estimated the 28-days pre-vaccination window separately since an event in that period may change the likelihood of receiving the vaccine.[44,45]

### Statistical analysis

We characterised the study participants in each cohort in terms of socio-demographics (e.g., age, sex), comorbidities (any time before vaccination), and recent medication use during the 6 months before the index date. We estimated the observed rates during the 28 days post-immunization and 90 days post-test period for the vaccinated and SARS-Cov-2 infected cohorts, respectively. Similarly, background rates were estimated for the general population in 2017– 2019, stratified by age and sex. We calculated crude incidence rates as the total number of events divided by the person-time at risk per 100,000 person-years. We used indirect standardisation to account for differences between the age-sex structure of the vaccinated or SARS-CoV-2 cohorts and the general population.[46] We calculated standardised incidence rate ratios (SIRs) and 95% confidence intervals (CIs) to compare observed and expected rates. The number needed to harm (NNH) were calculated for each cohort as well.[47]

We applied two further sensitivity analyses to check the impact of our design choices. We replicated the analyses after removing the 1-year prior observation requirement. We also recalculated background rates using a background population cohort of all participants followed from 1 January 2017 instead of using their first healthcare visit date as the index date.

In addition, a SCCS was conducted for any identified signals to minimise confounding by indication. Since individuals serve as their own controls, both measured and unmeasured time-fixed confounding are controlled by design. We included age (as 5-years band) and seasonality (four seasons) to adjust for time-varying confounding. Conditional Poisson regression models were fitted to estimate incidence rate ratio (IRR) and 95% confidence intervals for each outcome according to exposure window,[48] comparing post-vs pre-vaccine time. Due to the limited number of events, we only had power to run SCCS for Bell’s Palsy (Appendix table 2).[49] Any subgroups with less than 5 people were blinded and reported as <5, following information governance requirements. The study was approved by the Independent Scientific Advisory Committee (ISAC 20_000211). SCCS were run using the self-controlled case series package in R.[50] Our analytical code is available for review and replication of our findings at https://github.com/oxford-pharmacoepi/CovidVaccinationSafetyStudy_neuroimmune.

### Patient and public involvement

No patients or members of the public were directly involved in the design or analysis of the reported data. The Independent Scientific Advisory Committee responsible for the approval of our protocol involved patients in the evaluation of our data access application.

## RESULTS

We included 1.9 million people vaccinated with first-dose ChadOx1, 1.7 million vaccinated with first-dose BNT162b2, 299,311 infected with SARS-CoV-2, and 2.3 million general population participants. Both vaccinated cohorts were older (average age 66 and 67 years, respectively) than those infected with SARS-CoV-2 (average age 48 years) and the general population (average age 54 years). Vaccinated participants had a higher prevalence of comorbidities, including cancer, diabetes, and other long-term conditions comparing with the general population (see Table 1). The prevalence of use of systemic corticosteroids and anticoagulants/antithrombotics was also higher in the vaccinated cohorts than the infected cohort, but was similar to the general population used for estimating background rates.

**Table 1.** Baseline characteristics of the background population, those vaccinated with BNT162b2, those vaccinated with ChAdOx1, and those infected with SARS-CoV-2.

|  | **General population** | **Vaccinated with ChAdOx1** | **Vaccinated with BNT162b2** | **Infected with SARS-CoV-2** |
| --- | --- | --- | --- | --- |
| **N** | 2,290,537 | 1,868,767 | 1,661,139 | 299,311 |
| **Age (median [IQR])** | 54 [43 to 67] | 66 [56 to 73] | 67 [54 to 78] | 48 [39 to 58] |
| **Sex: Male** | 1,094,911 (47.8%) | 845,908 (45.3%) | 702,863 (42.3%) | 136,301 (45.5%) |
| **Years of prior observation time (median [IQR])** | 13.5 [8.1 to 16.3] | 19.0 [7.7 to 31.1] | 19.1 [7.8 to 31.2] | 12.4 [5.3 to 23.9] |
| ***Comorbidities*** |  |  |  |  |
| **Autoimmune disease** | 57,773 (2.5%) | 72,119 (3.9%) | 64,311 (3.9%) | 7,136 (2.4%) |
| **Malignancy** | 187,499 (8.2%) | 257,449 (13.8%) | 268,655 (16.2%) | 15,663 (5.2%) |
| **Diabetes mellitus** | 197,619 (8.6%) | 306,944 (16.4%) | 276,194 (16.6%) | 28,488 (9.5%) |
| **Obesity** | 94,054 (4.1%) | 120,052 (6.4%) | 98,900 (6.0%) | 16,791 (5.6%) |
| **Heart disease** | 251,708 (11.0%) | 334,469 (17.9%) | 349,669 (21.0%) | 24,511 (8.2%) |
| **Renal impairment** | 161,596 (7.1%) | 207,751 (11.1%) | 234,386 (14.1%) | 14,150 (4.7%) |
| ***Medication use (183 days prior to 4 days prior)*** |  |  |  |  |
| **Systemic corticosteroids** | 263,488 (11.5%) | 119,766 (6.4%) | 105,977 (6.4%) | 14,168 (4.7%) |
| **Antithrombotic and anticoagulant therapies** | 102,605 (4.5%) | 61,322 (3.3%) | 65,237 (3.9%) | 3,885 (1.3%) |

Among 1,867,708 ChAdOx1 vaccinees, we observed 64 cases of Bell’s palsy during the 28 days following vaccination and expected 48 events after standardisation, resulting in an SIR of 1.34 [1.05 to 1.72] (Table 2) and a number needed to harm (NNH) of 113,885. Equivalent figures for BNT162b2 were 54 observed and 47 expected among 1,660,289 participants, with a non-significant SIR of 1.15 [0.88 to 1.50]. Among 299,069 infected with SARS-CoV-2, we observed 21 events in the 90 days following a first positive PCR test and expected 17 events, equivalent to an SIR of 1.23 [0.80 to 1.89]. (Figure 2) The SCCS analysis showed adjusted IRR of 1.15 [0.87 to 1.49] for ChAdOx1, IRR of 1.10 [0.81 to 1.46] for BNT162b2, and IRR of 0.98 [0.61 to 1.52] for SARS-CoV-2 infection (Table 3).

**Table 2.** Association between COVID-19 vaccination or infection and the occurrence of neuro-immune adverse events of special interest.

| Outcome | Population | N | Person-years-at-risk | N observed | N expected | SIR | NNH |
| --- | --- | --- | --- | --- | --- | --- | --- |
| Bell's palsy | Vaccinated with ChAdOx1 | 1,867,708 | 119,078 | 64 | 47.6 | 1.34 (1.05 to 1.72) | 113885 |
|  | Vaccinated with BNT162b2 | 1,660,289 | 118,593 | 54 | 47 | 1.15 (0.88 to 1.50) | 237184 |
|  | SARS-CoV-2 PCR positive test | 299,069 | 40,573 | 21 | 17 | 1.23 (0.80 to 1.89) | 74767 |
| Encephalomyelitis | Vaccinated with ChAdOx1 | 1,868,554 | 119,134 | <5 | 2.5 | 1.62 (0.61 to 4.31) | 1245703 |
|  | Vaccinated with BNT162b2 | 1,661,019 | 118,646 | <5 | 2.3 | 0.86 (0.22 to 3.46) | -<br>5536730 |
|  | SARS-CoV-2 PCR positive test | 299,201 | 40,596 | 7 | 0.6 | 11.05 (5.27 to 23.17) | 46750 |
| Guillain-Barre syndrome | Vaccinated with ChAdOx1 | 1,868,598 | 119,136 | 6 | 3.1 | 1.91 (0.86 to 4.26) | 644344 |
|  | Vaccinated with BNT162b2 | 1,661,011 | 118,645 | <5 | 3.1 | 1.29 (0.49 to 3.45) | 1845568 |
|  | SARS-CoV-2 PCR positive test | 299,196 | 40,595 | <5 | 0.8 | 5.20 (1.95 to 13.85) | 93499 |
| Transverse myelitis | Vaccinated with ChAdOx1 | 1,868,613 | 119,138 | <5 | 1.3 | 2.27 (0.73 to 7.02) | 1099184 |
|  | Vaccinated with BNT162b2 | 1,661,035 | 118,647 | <5 | 1.3 | 2.26 (0.73 to 7.01) | 977079 |
|  | SARS-CoV-2 PCR positive test | 299,203 | 40,596 | <5 | 0.6 | 1.71 (0.24 to 12.15) | 748008 |

**SIR: standardised incidence ratio, NNH: number needed to harm**

**Table 3.** Incidence rate ratios (95% confidence intervals) for COVID-19 vaccination or infection and the occurrence of Bell’s Palsy.

| Time period | BNT162b2 |  | ChAdOx1 |  | SARS-CoV-2 PCR positive test |  |
| --- | --- | --- | --- | --- | --- | --- |
|  | Number of events | IRR | Number of events | IRR | Number of events | IRR |
| Baseline | 2918 | Ref | 3393 | Ref | 662 | Ref |
| -28 to -1 days pre-exposure | 57 | 1.13 (0.85 to 1.47) | 65 | 1.04 (0.80 to 1.34) | 14 | 1.10 (0.60 to 1.84) |
| 1-28 days post vaccination | 50 | 1.10 (0.81 to 1.46) | 58 | 1.15 (0.87 to 1.49) | - | - |
| 1-90 days post SARS-CoV-2 infection | - | - | - | - | 23 | 0.98 (0.61 to 1.52) |

**IRR: Incidence rate ratio**

Among 1,868,598 people vaccinated with ChAdOx1, 6 GBS events were observed and 3 expected, resulting in a non-significant SIR of 1.91 [0.86 to 4.26]. Among 1,661,011 people receiving BNT162b2, <5 GBS events were observed and 3 expected, resulting in an SIR of 1.29 [0.49 to 3.45]. In contrast, <5 GBS events were observed in 299,196 participants infected with SARS-CoV-2 and <1 expected, resulting in an SIR of 5.20 [1.95 to 13.85].

Sensitivity analyses were in line with these results. The main additional finding was a significant SIR for GBS following ChAdOx1 vaccination when we removed the 1-year washout and the requirement for a healthcare visit in the background population. We then observed 7 events in 1,868,598 participants and expected 3 events, resulting in an SIR of 2.20 [1.05 to 4.61], equivalent to an NNH of 514,773. No association was seen for BNT162b2. The sensitivity analysis had similar results to the main analysis for the cohort infected with SARS-CoV-2, with an SIR of 5.66 [2.12 to 15.07] and an NNH of 90,703. (Table 4)

**Table 4.**
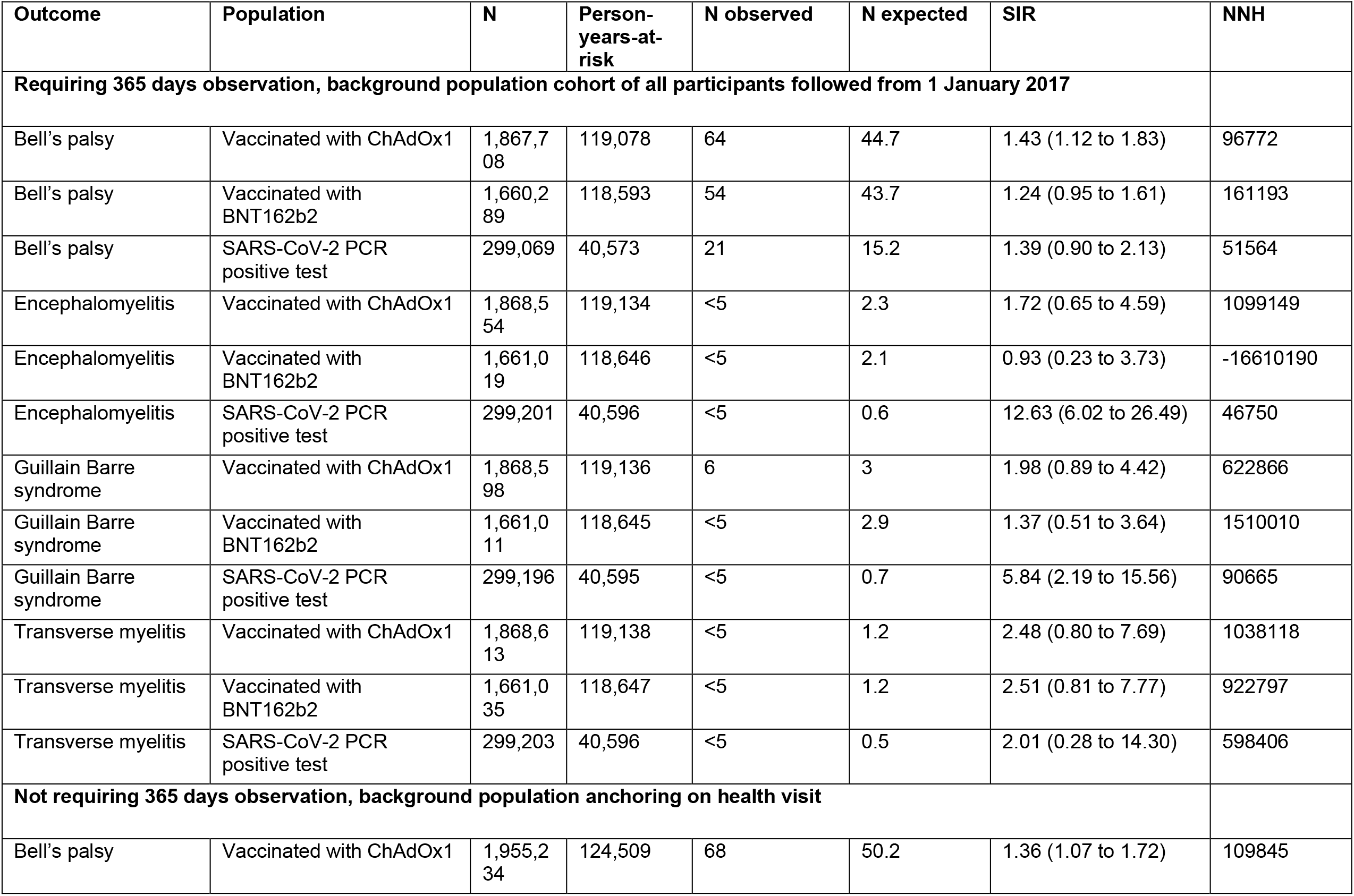
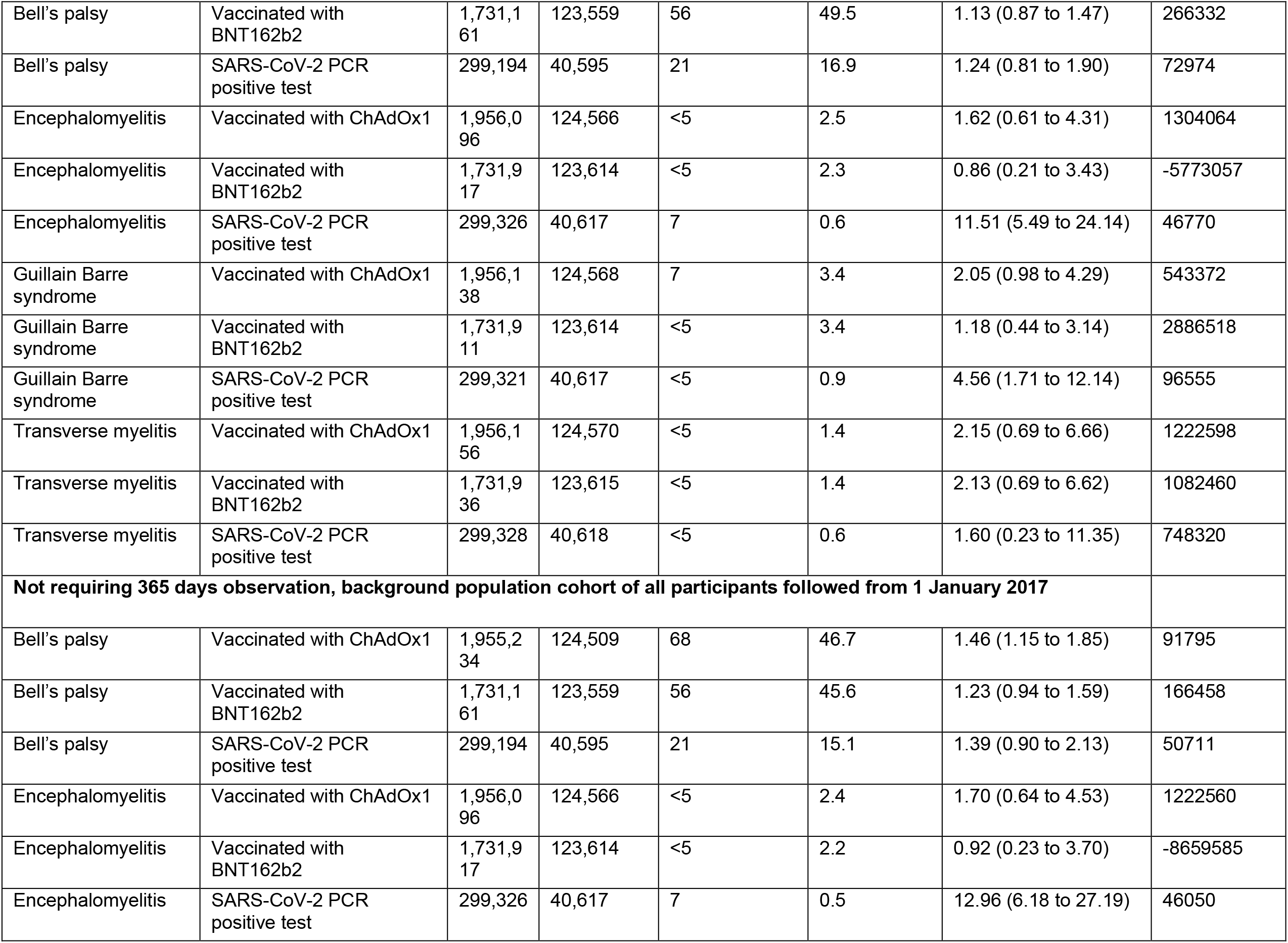
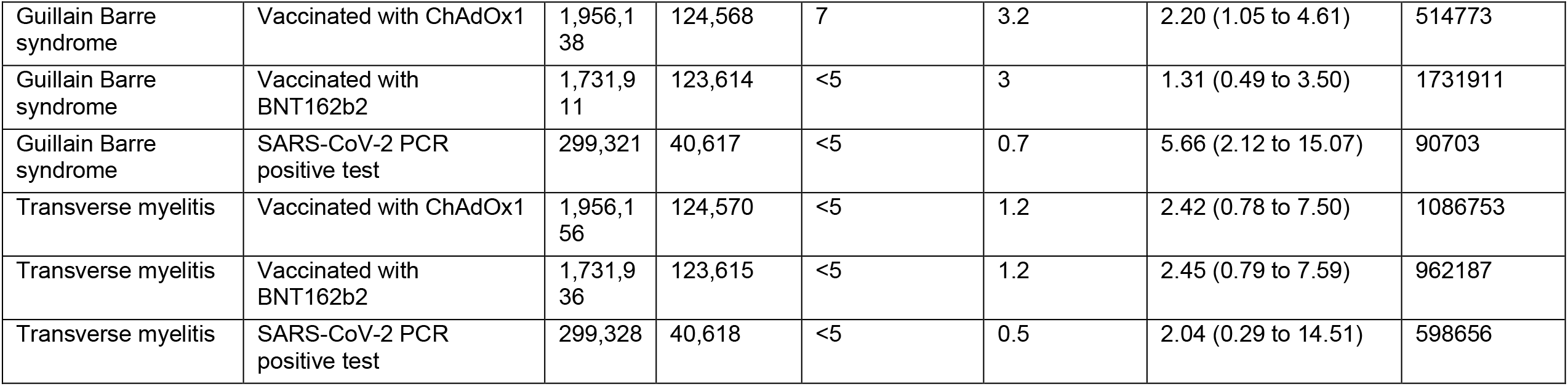
Sensitivity analyses.

| Outcome | Population | N | Person-years-at-risk | N observed | N expected | SIR | NNH |
| --- | --- | --- | --- | --- | --- | --- | --- |
| **Requiring 365 days observation, background population cohort of all participants followed from 1 January 2017** |  |  |  |  |  |  |  |
| Bell's palsy | Vaccinated with ChAdOx1 | 1,867,708 | 119,078 | 64 | 44.7 | 1.43 (1.12 to 1.83) | 96772 |
| Bell's palsy | Vaccinated with BNT162b2 | 1,660,289 | 118,593 | 54 | 43.7 | 1.24 (0.95 to 1.61) | 161193 |
| Bell's palsy | SARS-CoV-2 PCR positive test | 299,069 | 40,573 | 21 | 15.2 | 1.39 (0.90 to 2.13) | 51564 |
| Encephalomyelitis | Vaccinated with ChAdOx1 | 1,868,554 | 119,134 | <5 | 2.3 | 1.72 (0.65 to 4.59) | 1099149 |
| Encephalomyelitis | Vaccinated with BNT162b2 | 1,661,019 | 118,646 | <5 | 2.1 | 0.93 (0.23 to 3.73) | -16610190 |
| Encephalomyelitis | SARS-CoV-2 PCR positive test | 299,201 | 40,596 | <5 | 0.6 | 12.63 (6.02 to 26.49) | 46750 |
| Guillain Barre syndrome | Vaccinated with ChAdOx1 | 1,868,598 | 119,136 | 6 | 3 | 1.98 (0.89 to 4.42) | 622866 |
| Guillain Barre syndrome | Vaccinated with BNT162b2 | 1,661,011 | 118,645 | <5 | 2.9 | 1.37 (0.51 to 3.64) | 1510010 |
| Guillain Barre syndrome | SARS-CoV-2 PCR positive test | 299,196 | 40,595 | <5 | 0.7 | 5.84 (2.19 to 15.56) | 90665 |
| Transverse myelitis | Vaccinated with ChAdOx1 | 1,868,613 | 119,138 | <5 | 1.2 | 2.48 (0.80 to 7.69) | 1038118 |
| Transverse myelitis | Vaccinated with BNT162b2 | 1,661,035 | 118,647 | <5 | 1.2 | 2.51 (0.81 to 7.77) | 922797 |
| Transverse myelitis | SARS-CoV-2 PCR positive test | 299,203 | 40,596 | <5 | 0.5 | 2.01 (0.28 to 14.30) | 598406 |
| **Not requiring 365 days observation, background population anchoring on health visit** |  |  |  |  |  |  |  |
| Bell's palsy | Vaccinated with ChAdOx1 | 1,955,234 | 124,509 | 68 | 50.2 | 1.36 (1.07 to 1.72) | 109845 |
| Bell's palsy | Vaccinated with BNT162b2 | 1,731,161 | 123,559 | 56 | 49.5 | 1.13 (0.87 to 1.47) | 266332 |
| Bell's palsy | SARS-CoV-2 PCR positive test | 299,194 | 40,595 | 21 | 16.9 | 1.24 (0.81 to 1.90) | 72974 |
| Encephalomyelitis | Vaccinated with ChAdOx1 | 1,956,096 | 124,566 | <5 | 2.5 | 1.62 (0.61 to 4.31) | 1304064 |
| Encephalomyelitis | Vaccinated with BNT162b2 | 1,731,917 | 123,614 | <5 | 2.3 | 0.86 (0.21 to 3.43) | -5773057 |
| Encephalomyelitis | SARS-CoV-2 PCR positive test | 299,326 | 40,617 | 7 | 0.6 | 11.51 (5.49 to 24.14) | 46770 |
| Guillain Barre syndrome | Vaccinated with ChAdOx1 | 1,956,138 | 124,568 | 7 | 3.4 | 2.05 (0.98 to 4.29) | 543372 |
| Guillain Barre syndrome | Vaccinated with BNT162b2 | 1,731,911 | 123,614 | <5 | 3.4 | 1.18 (0.44 to 3.14) | 2886518 |
| Guillain Barre syndrome | SARS-CoV-2 PCR positive test | 299,321 | 40,617 | <5 | 0.9 | 4.56 (1.71 to 12.14) | 96555 |
| Transverse myelitis | Vaccinated with ChAdOx1 | 1,956,156 | 124,570 | <5 | 1.4 | 2.15 (0.69 to 6.66) | 1222598 |
| Transverse myelitis | Vaccinated with BNT162b2 | 1,731,936 | 123,615 | <5 | 1.4 | 2.13 (0.69 to 6.62) | 1082460 |
| Transverse myelitis | SARS-CoV-2 PCR positive test | 299,328 | 40,618 | <5 | 0.6 | 1.60 (0.23 to 11.35) | 748320 |
| **Not requiring 365 days observation, background population cohort of all participants followed from 1 January 2017** |  |  |  |  |  |  |  |
| Bell's palsy | Vaccinated with ChAdOx1 | 1,955,234 | 124,509 | 68 | 46.7 | 1.46 (1.15 to 1.85) | 91795 |
| Bell's palsy | Vaccinated with BNT162b2 | 1,731,161 | 123,559 | 56 | 45.6 | 1.23 (0.94 to 1.59) | 166458 |
| Bell's palsy | SARS-CoV-2 PCR positive test | 299,194 | 40,595 | 21 | 15.1 | 1.39 (0.90 to 2.13) | 50711 |
| Encephalomyelitis | Vaccinated with ChAdOx1 | 1,956,096 | 124,566 | <5 | 2.4 | 1.70 (0.64 to 4.53) | 1222560 |
| Encephalomyelitis | Vaccinated with BNT162b2 | 1,731,917 | 123,614 | <5 | 2.2 | 0.92 (0.23 to 3.70) | -8659585 |
| Encephalomyelitis | SARS-CoV-2 PCR positive test | 299,326 | 40,617 | 7 | 0.5 | 12.96 (6.18 to 27.19) | 46050 |
| Guillain Barre syndrome | Vaccinated with ChAdOx1 | 1,956,138 | 124,568 | 7 | 3.2 | 2.20 (1.05 to 4.61) | 514773 |
| Guillain Barre syndrome | Vaccinated with BNT162b2 | 1,731,911 | 123,614 | <5 | 3 | 1.31 (0.49 to 3.50) | 1731911 |
| Guillain Barre syndrome | SARS-CoV-2 PCR positive test | 299,321 | 40,617 | <5 | 0.7 | 5.66 (2.12 to 15.07) | 90703 |
| Transverse myelitis | Vaccinated with ChAdOx1 | 1,956,156 | 124,570 | <5 | 1.2 | 2.42 (0.78 to 7.50) | 1086753 |
| Transverse myelitis | Vaccinated with BNT162b2 | 1,731,936 | 123,615 | <5 | 1.2 | 2.45 (0.79 to 7.59) | 962187 |
| Transverse myelitis | SARS-CoV-2 PCR positive test | 299,328 | 40,618 | <5 | 0.5 | 2.04 (0.29 to 14.51) | 598656 |

Fewer than 5 events of encephalomyelitis were seen following ChAdOx1 and BNT162b2, with equivalent SIRs of 1.62 [0.61 to 4.31] and 0.86 [0.22 to 3.46], respectively. Seven encephalomyelitis events were observed and <1 expected following SARS-CoV-2 infection in 299,201 participants, equating to an SIR of 11.05 [5.27 to 23.17], and an NNH of 46,750. All sensitivity analyses were in line with these results.

Transverse myelitis was very rare in all three cohorts, with <5 events observed and expected in the vaccinated and infected cohorts. Equivalent adjusted SIRs were 2.27 [0.73 to 7.02] in the ChAdOx1 cohort, 2.26 [0.73 to 7.01] in the BNT162b2 cohort, and 1.71 [0.24 to 12.15] in the infected cohort. Sensitivity analyses were consistent with these results.

## DISCUSSION

### Key findings

This is the largest cohort study to date on the neuro-immune safety of COVID-19 vaccines, including 1.9 million ChAdOx1 and 1.7 million BNT162b2 vaccine recipients. Our primary analyses with a historical rates comparison method found a potential association between ChAdOx1 and the risk of developing Bell’s palsy, with an estimated 30% relative increase in risk. If causal, this signal would equate to one Bell’s palsy event per 110,000 people receiving this vaccine. No associations were found with the BNT162b2 vaccine or SARS-CoV-2 infection. A SCCS to minimise patient-level confounding showed no statistically significant association between either vaccine and Bell’s palsy. As we observed differences in baseline characteristics between vaccine recipients and the background population, our findings could be related to residual confounding rather than a causal link.

We analysed a cohort of almost 300,000 people infected with the virus these vaccines prevent, SARS-CoV-2. We demonstrated a striking association between COVID-19 and two severe immune-mediated neurological outcomes: a 5-fold increased risk of GBS and an 11-fold excess risk of encephalomyelitis. These findings provide a useful counterfactual for vaccination and strengthen the risk-benefit of achieving immunity against COVID-19 through vaccination rather than infection.

### Findings in context

Neuro-immune disorders have been identified as adverse events of special interest by regulators, such as the FDA in the US and EMA in Europe, and organisations such as the Brighton Collaboration. They have been closely monitored during immunisation campaigns.[43,51,52] Although phase 3 clinical trials of mRNA vaccines noted an imbalance of Bell’s palsy cases in vaccine and placebo groups, there was insufficient power to draw causal associations.[2,53]

More recently, case reports and case series have reported immune-mediated neurological disorders after COVID-19 vaccines, such as facial nerve palsies after mRNA Covid-19 vaccines[54] and cases of GBS after adenovirus-based vaccines, including ChAdOx1.[7–9,13] On 13 July 2021, the FDA updated the label of the Johnson & Johnson adenovirus-based COVID-19 vaccine to include a warning about the potential risk of GBS following vaccination.[55]

The product information for the ChAdOx1 vaccine has also included a warning that Guillain-Barré syndrome (GBS) has very rarely followed vaccination.[37] However, there is currently not enough data on the safety of vaccines for people with these conditions. Conversely, the UK MHRA stated on 16 July 2021 that the number of reports of Bell’s palsy from the spontaneous report system is similar to the expected rate and that there is no increased risk following any vaccine based on current evidence.[6]

Real-world studies have also assessed the association between facial nerve palsy, SARS-CoV-2 infection and COVID-19 vaccines. One study using electronic medical records data from 41 healthcare organisations worldwide found that 0.08% of patients had a diagnosis of Bell’s palsy within 8 weeks after being diagnosed with COVID-19, of whom 53.9% had no history of Bell’s palsy, and a higher risk in patients with COVID-19 compared with those vaccinated. However, they did not calculate the comparative risk of Bell’s palsy between COVID-19 vaccination and the general population.[56] A case-control study from Israel analysed emergency department admission data and found no increased risk of facial nerve palsy after BNT162b2 vaccination. They also compared the numbers of cases in January and February 2021 to the same period during 2015 to 2020, and found no meaningful increase in admission numbers due to facial nerve palsy.[57] Renoud et al. conducted a disproportionality analysis using the World Health Organization’s pharmacovigilance database. They found no higher safety signal for facial palsy after mRNA Covid-19 vaccines when compared with all other viral vaccines or restricted to influenza vaccines.[58] Eric Wan et al. found an overall increased risk of Bell’s palsy after vaccinated with CoronaVac, an inactivated virus COVID-19 vaccine, but no increased risk after BNT162b2 vaccination.[59] In our study, we found no elevated risk of Bell’s palsy with BNT162b2, but a small increased risk with ChAdOx1. Yet the association was not proved in the SCCS analysis where confounding was addressed.

Although cases of neuro-immune disorders after SARS-CoV-2 infection have been reported,[60–63] causality remains unclear. Single cases of GBS and small series, mainly from Europe and including all phenotypic variants, were reported and postulated a possible post-infectious mechanism.[64–66] Multicentric studies in Spain and Italy during the beginning of the pandemic reported a higher frequency of GBS among patients with COVID-19 than those without COVID-19, or compared with the incidence in the same months of 2019.[67,68] However, one study compared reported cases in 2020 and 2016–2019 in the England National Immunoglobulin Database and found no significant association between COVID-19 and GBS.[69] Another study reported a 7-fold risk of Bell’s palsy among patients with COVID-19 compared with individuals who received COVID-19 vaccines.[56] Recent studies have evaluated reports of acute disseminated encephalomyelitis and acute haemorrhagic leukoencephalitis and identified more than 45 cases up to May-June, 2021 associated with SARS-CoV-2 infection and with poor neurologic outcomes.[70,71] Our study found an 11-fold increased risk for encephalomyelitis and 5-fold for GBS after SARS-CoV-2 infection, with no elevated risk for Bell’s palsy or transverse myelitis.

### Study strengths and limitations

The main strength of our study is the population-based method with data from the UK primary care system, which have great representativeness and complete capture of vaccinations among the UK population. In addition, the use of self-controlled case series study design took the advantage of within person comparison to reduce time-fixed confounding. We also further adjusted for age and seasonality to control for time-varying confounding.

Our study has limitations. As we only included primary care data, diagnoses from inpatient settings may not be captured and the absolute risk may be underestimated. However, a previous study has showed that CPRD primary care data captures immune-mediated neurological disorders like GBS accurately, even without linking hospital data.[72] Within-database comparisons are recommended when comparing observed and expected rates due to database-level heterogeneity.[43] We used CPRD Aurum and CPRD GOLD. Although both contain UK primary care data, these data come from different electronic health record clinical systems and use different medical coding. However, one study showed that the two databases generated similar estimates.[73] We used code lists and algorithms for the identification of neuro-immune events previously published as part of a study on the background rates of COVID-19 AESI.[43] Mapping to the OMOP common data model also helped maximise comparability. The short follow-up time after vaccination may have led to underreporting of events of interest.

The background rate comparison method has been shown to be a sensitive but not specific method, with a high type 1 error.[74] Although we adjusted for age and sex by indirect standardisation and anchored the background incidence rates on a health visit or contact as recommended, the analysis may still have overestimated the risk. Our SCCS analyses provide further reassurance, as they account for all time-fixed confounding by using the same person as a “control”.[44]

## Conclusion

We found no consistent risk of any of the studied neuro-immune events with either vaccine. Conversely, we found a 5-fold increase in risk of GBS and an 11-fold risk of encephalomyelitis following COVID-19 infection. Our data illustrate that SARS-CoV-2 infection, and not COVID-19 vaccines, is associated with an increased risk of neuro-immune outcomes.

## Data Availability

Patient level data cannot be shared without approval from data custodians owing to local information governance and data protection regulations. Aggregated data, analytical code, and detailed definitions of algorithms for identifying the events are available in a GitHub repository (https://github.com/oxford-pharmacoepi/CovidVaccinationSafetyStudy_neuroimmune).

## Ethical approval

The protocol for this research was approved by the independent scientific advisory committee for Medicine and Healthcare products Regulatory Agency database research (protocol number 20_000211)

## Acknowledgments

We thank Jennifer A de Beyer (Centre for Statistics in Medicine, University of Oxford) for English language editing.

## Contributors

XL and EMH are joint first authors. XL, DAP, EB and VS conceived the study and contributed to the study design. XL, VS, and DAP interpreted the results and wrote the manuscript. All coauthors contributed to writing the manuscript. All authors approved the final version and had final responsibility for the decision to submit for publication.

## Competing interests

All authors have completed the ICMJE disclosure form at http://www.icmje.org/disclosure-of-interest/ and declare the following interests: DPA receives funding from the UK National Institute for Health Research (NIHR) in the form of a senior research fellowship and the Oxford NIHR Biomedical Research Centre. XL receives the Clarendon Fund and Brasenose College Scholarship (University of Oxford) to support her DPhil study. DPA’s research group has received research grants from the European Medicines Agency; the Innovative Medicines Initiative; Amgen, Chiesi, and UCB Biopharma; and consultancy or speaker fees from Astellas, Amgen, and UCB Biopharma.

The lead authors (XL and EMH) affirm that this manuscript is an honest, accurate, and transparent account of the study being reported; that no important aspects of the study have been omitted; and that any discrepancies from the study as planned (and, if relevant, registered) have been explained.

Dissemination to participants and related patient and public communities: We will disseminate a lay summary of our findings through our Twitter and other social media accounts.

## APPENDIX

**Table 1:** Medical codes for outcomes.

| Id | Code | Name | Vocabulary |
| --- | --- | --- | --- |
| **Bell's palsy** |  |  |  |
| 44784628 | 87651000119100 | Facial palsy House-Brackmann grade VI | SNOMED |
| 44782796 | 68151000119106 | Facial palsy House-Brackmann grade V | SNOMED |
| 44782795 | 68141000119109 | Facial palsy House-Brackmann grade IV | SNOMED |
| 44782794 | 68131000119100 | Facial palsy House-Brackmann grade III | SNOMED |
| 44782526 | 68121000119103 | Facial palsy House-Brackmann grade II | SNOMED |
| 42710044 | 449916008 | Peripheral facial palsy | SNOMED |
| 37396434 | 716243005 | Sellars Beighton syndrome | SNOMED |
| 37204553 | 783257005 | Familial recurrent peripheral facial palsy | SNOMED |
| 4091559 | 280816001 | Facial palsy | SNOMED |
| 4063208 | 164110008 | O/E - cranial 7 -paralysis-UMN | SNOMED |
| 4044386 | 230546006 | Familial facial nerve palsy | SNOMED |
| 4002030 | 11045000 | Supranuclear facial nerve paralysis | SNOMED |
| 3182576 | 12870001000004100 | Partial seventh nerve palsy | Nebraska Lexicon |
| 760893 | 12239661000119100 | Bells palsy of right side of face | SNOMED |
| 760892 | 12239621000119100 | Bells palsy of left side of face | SNOMED |
| 374923 | 193093009 | Bell's palsy | SNOMED |
| **transverse myelitis** |  |  |  |
| 4047628 | 230197009 | Acute viral transverse myelitis | SNOMED |
| 4044077 | 230378004 | Acute non-infective transverse myelitis | SNOMED |
| 4042204 | 230198004 | Varicella transverse myelitis | SNOMED |
| 443904 | 16631009 | Transverse myelopathy syndrome | SNOMED |
| 139803 | 47000000 | Acute transverse myelitis | SNOMED |
| 134330 | 425756000 | Idiopathic transverse myelitis | SNOMED |
| **Guillain-Barré syndrome** |  |  |  |
| 37396786 | 716723000 | Acute inflammatory demyelinating polyradiculoneuropathy | SNOMED |
| 37396785 | 716722005 | Acute motor sensory axonal neuropathy | SNOMED |
| 37396086 | 715770009 | Acute motor axonal neuropathy | SNOMED |
| 37204541 | 783244002 | Acute pure sensory neuropathy | SNOMED |
| 37204369 | 783010003 | Pharyngeal-cervical-brachial variant of Guillain-Barré syndrome | SNOMED |
| 35624130 | 766722008 | Paraparetic variant of Guillain-Barré syndrome | SNOMED |
| 35623634 | 766049000 | Acute sensory ataxic neuropathy | SNOMED |
| 4164770 | 40956001 | Guillain-Barré syndrome | SNOMED |
| 4070552 | 1767005 | Fisher's syndrome | SNOMED |
| 374925 | 129131007 | Acute infective polyneuritis | SNOMED |
| **encephalomyelitis** |  |  |  |
| 46285143 | 963741000000101 | Bacterial meningoencephalomyelitis | SNOMED |
| 43530701 | 182961000119101 | Acute disseminated encephalomyelitis following infectious disease | SNOMED |
| 40480147 | 443153007 | Toxic encephalomyelitis | SNOMED |
| 36715541 | 721249007 | Infection causing encephalomyelitis | SNOMED |
| 36676712 | 773688007 | Chronic lymphocytic inflammation with pontine perivascular enhancement responsive to steroids | SNOMED |
| 35607947 | 763310000 | Acute necrotizing encephalopathy of childhood | SNOMED |
| 4330496 | 431520004 | Inflammation of spinal cord due to toxin | SNOMED |
| 4320933 | 70576008 | Enteroviral encephalomyelitis | SNOMED |
| 4310021 | 424761009 | Paraneoplastic encephalomyelitis | SNOMED |
| 4249574 | 72986009 | Acute hemorrhagic leukoencephalitis | SNOMED |
| 4242575 | 58766009 | Granulomatous meningoencephalomyelitis | SNOMED |
| 4233538 | 359686005 | Van Bogaert's sclerosing leukoencephalitis | SNOMED |
| 4228612 | 422089004 | Encephalomyelitis associated with AIDS | SNOMED |
| 4220014 | 397961002 | Encephalomyelitis caused by lymphocytic choriomeningitis virus | SNOMED |
| 4206037 | 55234004 | Pyogranulomatous meningoencephalomyelitis | SNOMED |
| 4190307 | 39367000 | Inflammatory disease of the central nervous system | SNOMED |
| 4176250 | 50776006 | Allergic encephalomyelitis | SNOMED |
| 4147498 | 267576008 | Encephalitis, myelitis and encephalomyelitis | SNOMED |
| 4120303 | 302811004 | Progressive congenital rubella encephalomyelitis | SNOMED |
| 4105201 | 192724008 | Post mixed vaccination encephalitis | SNOMED |
| 4105200 | 192711008 | Post diphtheria vaccination encephalitis | SNOMED |
| 4104690 | 192723002 | Post hepatitis B vaccination encephalitis | SNOMED |
| 4104546 | 29197009 | Meningoencephalomyelitis | SNOMED |
| 4103116 | 192721000 | Post influenza vaccination encephalitis | SNOMED |
| 4103115 | 192719005 | Post mumps vaccination encephalitis | SNOMED |
| 4103114 | 192718002 | Post polio vaccination encephalitis | SNOMED |
| 4103113 | 192716003 | Post yellow fever vaccination encephalitis | SNOMED |
| 4103112 | 192714000 | Post rabies vaccination encephalitis | SNOMED |
| 4103111 | 192712001 | Post pertussis vaccination encephalitis | SNOMED |
| 4103110 | 192709004 | Post plague vaccination encephalitis | SNOMED |
| 4103109 | 192708007 | Post cholera vaccination encephalitis | SNOMED |
| 4103108 | 192707002 | Post paratyphoid vaccination encephalitis | SNOMED |
| 4103107 | 192706006 | Post typhoid vaccination encephalitis | SNOMED |
| 4102199 | 192717007 | Post measles vaccination encephalitis | SNOMED |
| 4102198 | 192705005 | Post BCG vaccination encephalitis | SNOMED |
| 4100112 | 192722007 | Post hepatitis A vaccination encephalitis | SNOMED |
| 4100111 | 192720004 | Post rubella vaccination encephalitis | SNOMED |
| 4100110 | 192715004 | Post typhus vaccination encephalitis | SNOMED |
| 4100109 | 192713006 | Post smallpox vaccination encephalitis | SNOMED |
| 4100108 | 192710009 | Post tetanus vaccination encephalitis | SNOMED |
| 4098889 | 27040004 | Experimental allergic encephalomyelitis | SNOMED |
| 4042202 | 230188005 | Post-influenza encephalitis | SNOMED |
| 4035637 | 15316002 | Simian B encephalomyelitis | SNOMED |
| 3655306 | 860842004 | Encephalomyelitis caused by Coxiella burnetii | SNOMED |
| 3655305 | 860841006 | Encephalomyelitis caused by bacterium | SNOMED |
| 3655275 | 860805006 | Encephalomyelitis caused by Neisseria meningitidis | SNOMED |
| 3654623 | 838315004 | Encephalomyelitis caused by Burkholderia | SNOMED |
| 765827 | 432451000124103 | Multiphasic acute disseminated encephalomyelitis | SNOMED |
| 761989 | 2701000124105 | Recurrent acute disseminated encephalomyelitis | SNOMED |
| 443802 | 31367003 | Postvaccinal encephalomyelitis | SNOMED |
| 380322 | 186509002 | Postvaricella encephalitis | SNOMED |
| 379798 | 230199007 | Post-infectious encephalomyelitis | SNOMED |
| 379792 | 192704009 | Post-immunization encephalitis | SNOMED |
| 378143 | 45170000 | Encephalitis | SNOMED |
| 377487 | 111872008 | Post measles encephalitis | SNOMED |
| 375185 | 128191000 | Rubella encephalomyelitis | SNOMED |
| 374021 | 83942000 | Acute disseminated encephalomyelitis | SNOMED |
| 373189 | 62950007 | Encephalomyelitis | SNOMED |
| 372615 | 192727001 | Post-infectious encephalitis | SNOMED |
| 138965 | 41370002 | Myelitis | SNOMED |

**Table 2:** Power calculation for SCCS: minimum detectable relative risk ChAdOx1.

**ChAdOx1**
| **Outcome** | **Proportion time exposed** | **Proportion population exposed** | **Events** | **MDRR** |
| --- | --- | --- | --- | --- |
| **Bell's palsy** | 0.0346 | 0.9883 | 3456 | 1.2791 |
| **Encephalomyelitis** | 0.0346 | 0.9883 | 251 | 2.2298 |
| **Guillain Barre syndrome** | 0.0346 | 0.9883 | 206 | 2.3894 |
| **Transverse myelitis** | 0.0346 | 0.9883 | 147 | 2.7197 |

**BNT162b2**
| **Outcome** | **Proportion time exposed** | **Proportion population exposed** | **Events** | **MDRR** |
| --- | --- | --- | --- | --- |
| **Bell's palsy** | 0.0347 | 0.9899 | 3485 | 1.2777 |
| **Encephalomyelitis** | 0.0347 | 0.9899 | 198 | 2.4230 |
| **Guillain Barre syndrome** | 0.0347 | 0.9899 | 253 | 2.2229 |
| **Transverse myelitis** | 0.0347 | 0.9899 | 98 | 3.2415 |

**MDRR:** minimal detectable relative risk

